# Trajectories of physical activity after ischemic stroke: exploring prediction of change

**DOI:** 10.1101/2024.07.01.24309788

**Authors:** Andreas Gammelgaard Damsbo, Rolf Ankerlund Blauenfeldt, Grethe Andersen, Søren P. Johnsen, Janne Kaergaard Mortensen

## Abstract

**Background and aims:** Physical activity (PA) is associated with lower risk of stroke and better functional outcome. However, sedentary behavior after stroke is prevalent. We aimed to identify predictors for PA after first-time ischemic stroke and to develop prediction models to assess change in PA after stroke.

**Methods:** We used the Physical Activity Scale for the Elderly (PASE) to quantify PA prior to stroke and six months after stroke. Demographic and clinical data were enriched with registry data on socioeconomic status (SES). Associations with post-stroke PA were analyzed using linear regression. Elastic net regression models were used to predict decrease from higher PASE quartile to the lowest and increase from lowest to higher.

**Results:** A total of 523 patients, from The Efficacy of Citalopram Treatment in Acute Ischemic Stroke (TALOS) trial, had complete PASE data. Median [IQR] age was 69 years [59, 77], 181 (35%) were female and median [IQR] National Institute of Health Stroke Scale score was 3 [2, 5]. Overall, median PASE score did not change, but 20 % of patients decreased to the lowest PASE quartile whereas 48% from the lowest PASE quartile increased to a higher level. The prediction model performance, as represented by the area under receiver operating curve, was 0.679 for predicting a decrease and 0.619 for predicting an increase in PA. SES was the most consistent predictor for both decrease and for a lack of increase in PA.

**Conclusions:** Almost 1/2 of the least active patients increased their PA after stroke whereas 1/5 decreased to the least active level with socioeconomic factors being the most consistent predictors for change. Despite including comprehensive data, the PA prediction models had a modest predictive performance. Efforts to optimize PA should include all stroke survivors to improve PA for previously inactive patients and prevent PA decrease.

## Introduction

Within the past three decades, stroke outcome has improved as a result of optimized primary prophylaxis, acute stroke treatment and rehabilitation.^1–4^ Although survival has improved, life after stroke may still be complicated by depression, cognitive impairment, physical impairment, and lower quality of life.^5,6^

In the 1950s, a series of papers were published to investigate the beneficial effects of physical activity (PA) on cardio-vascular risk factors for the first time.^7,8^ Since then the body of evidence for the positive effect of PA on stroke risk has been growing.^9^ Studies have shown that patients with a higher pre-stroke PA level, experience less acute infarct growth^10^, fewer depressive symptoms,^11^ and better cognitive outcome.^12,13^ Studies have shown that any increase in PA is clinically relevant, and that sedentary behavior should be avoided, which has also been included in recent guidelines.^14–17^

Despite the mounting evidence of PA as an important modifiable risk factor, PA intervention after stroke has proven difficult.^18–20^ The level of PA is likely to be affected by stroke, due to motor impairment and fear of falling, and depression or cognitive impairment may decrease PA.^21,22^ Inequality in health driven by socioeconomic status may also be important as low socioeconomic status affects the level of PA among the elderly in general,^23^ affects the chance of receiving reperfusion treatment and is associated to lower functional outcome.^24^

The aim of this study was to explore changes from pre-stroke to post-stroke PA. Further, we aimed to develop a prediction tool based on socioeconomical data and other available variables at stroke onset that may identify patients with a risk of decrease and patients likely to increase PA post-stroke.

## Methods

This study is a post-hoc analysis of data from The Efficacy of Citalopram Treatment in Acute Ischemic Stroke (TALOS) trial, which was a randomized, double-blinded, placebo-controlled multicenter study on citalopram treatment after ischemic stroke. The trial inclusion period was from 2013 to 2016.^25^

Inclusion and exclusion criteria have been reported in detail elsewhere.^26^ In short, the study included patients with a first time ischemic stroke within the past seven days. Patients not able to give consent were included with consent from next of kin, and patients with a diagnosis of dementia or other neurodegenerative disease, antidepressant treatment, indications of noncompliant behavior, other life-threatening diseases, or contraindications to citalopram were not included.^26^ Patients without data on PA were excluded from the present study (n=119, 19%).

Individual-level clinical data may be shared upon reasonable request after finalizing a collaboration agreement. Registry-based data is available from relevant public registries after completing registration and relevant applications.

### Primary outcome

Physical activity was assessed using the Physical Activity Scale in the Elderly (PASE) questionnaire. The PASE questionnaire is designed to incorporate both exercise as well as more general activities including housework, gardening, caring for others and time spent at work. The PASE is a validated 12-item self-assessed questionnaire on overall PA during the past seven days. The score ranges from 0 to 793 with a higher score corresponding to a higher level of PA.^27^ Information on pre-stroke PA was obtained at inclusion where the patients were asked to report on the level of PA during the past seven days prior to stroke onset. The same questionnaire was used at six months follow-up.

The PASE score was categorized into quartiles based on pre-stroke PASE score. The full dataset was split based on pre-stroke PASE score into quartile 2-4 and quartile 1 for developing prediction models. The outcomes for the two models respectively, was defined as a decrease to the lowest PASE score quartile at six months or increase to any higher PASE score quartile at six months.

### Potential predictors

#### Clinical data

Baseline data was obtained from the Danish Stroke Registry.^28^ This registry contains detailed information on stroke patients in Denmark with a high level of completeness.^29^ These data include information on stroke severity at admission, measured by the National Institute of Health Stroke Scale (NIHSS) score, sex, thrombolysis and/or thrombectomy treatment, living arrangements, tobacco smoking, alcohol consumption (defined as above 7/14 units daily for females/males), hypertension, diabetes, prior transitory ischemic attack, newly diagnosed or known atrial fibrillation, prior myocardial infarction, peripheral arterial disease and body mass index (BMI).

Well-being was assessed at admission using the World Health Organization 5-item Well-being Index (WHO-5) for a continuous score of 0-100 range, with higher scores indicating better well-being.^30^ Functional status was recorded using the categorical modified Rankin Scale (mRS). The score ranges from 0 to 6, with higher scores indicating a greater deficit. The score was dichotomized for a score of 0 or > 0.^31^

No data on ethnicity was included.

#### Socioeconomic status

Socioeconomic data was retrieved from relevant registries at Statistics Denmark.^32^ Information on income, education and employment status were included to assess socioeconomic status (SES). All were defined at the time of study inclusion. We considered income group as the overall tertile (low, medium, high) of family equivalent disposable (FED) income averaged over the five calendar years preceding index stroke. The FED income accounts for members in the household as well as the rental value of property owned. Education was defined as the highest attained educational level, categorized according to The International Standard Classification of Education and divided into three categories: low (0-2), medium (3-4) and high (5-9).^33^ Employment status was defined according to the International Classification of Status in Employment. The classification was dichotomized in working or not working (not employed or retired) as only few very were not employed and not retired.

All data was linked on the individual level based on the Danish Civil Registration Number. Due to data safety regulations governing registry-based data, counts below five are rounded.

### Statistics

Pre-stroke and six months post-stroke PASE scores were compared using a paired Wilcoxon rank sum test of different medians.

Baseline characteristics of patients are shown stratified by category of change in PA level. Changes in PASE score quartiles are visualized with a Sankey plot.

A complete dataset was created using chained equations (predictive mean matching) for continuous variables and logistic regression for binary. A total of 10 imputations with each 10 iterations were generated. Regression models were created for each imputed dataset and results were pooled across all models.

#### Regression analysis

Univariable and multivariable linear regression models were created with follow-up PASE score as the outcome. Results are presented as mean difference in PASE score with 95 % confidence intervals (CI). Analyses were performed using the raw data set excluding body mass index due to a high proportion of patients with missing values and repeated using a complete dataset after multiple imputation for multivariable analysis.

#### Prediction models

The prediction models were based on elastic net logistic regression for outcome classification.^34^ All variables were standardized and coefficients were returned on original scale. The prediction models were tuned with regards to penalty ratio (alpha-values, 0 to 1 by 0.1 increments) through nested 10-fold cross validation with selection of model parameters for the model with lowest mean squared error. The final model is 10-fold cross validated. Prediction models were based on the complete dataset after imputation with results pooled across all imputed datasets.

This elastic net model type was chosen, as it provides model coefficients for inference. No CIs are available, however. Coefficients are presented as both median and mean odds ratios (OR) based on cross validation. Variables with both median and mean OR ≠ 1 are reported as significant. Median coefficients are visualized in a bar plot. The five most important variables (highest absolute log(OR)) for each model is reported in the text.

Predictive performance measures include sensitivity, specificity, positive predictive value, negative predictive value and area under receiver operated curve (AUROC) as well as a calibration plot. For clinical application, the goal was an AUROC of at least 0.7.

### Software

All analyses were performed using R Statistical Software 4.3.3 and all packages were installed from The Comprehensive R Archive Network.^35^ Plots were created with ggplot2 3.5.0 and ggalluvial 0.12.5.^36,37^ Prediction models were developed using glmnet 4.1-8.^38^ Multiple imputation was performed using mice 3.16.0.^39^

## Results

Overall, 523 patients had PASE scores available at both baseline and six months. Median [Interquartile range, IQR] pre-stroke PASE score was 137 [84; 202] with a total range of 0 to 574. At six months, the median PASE score was 135 [78; 206] with a range of 0 to 486. These were not statistically different. Median [IQR] age was 69 [59, 77], ranging from 19 to 99 and 181 (35%) were females. Median [IQR] NIHSS score was 3 [2, 5] and 197 patients (38 %) received either intravenous thrombolysis, endovascular treatment or both. See Table 1 for more details. Demographic and clinical data on the complete TALOS population has been reported previously.^25^

**Table 1:**
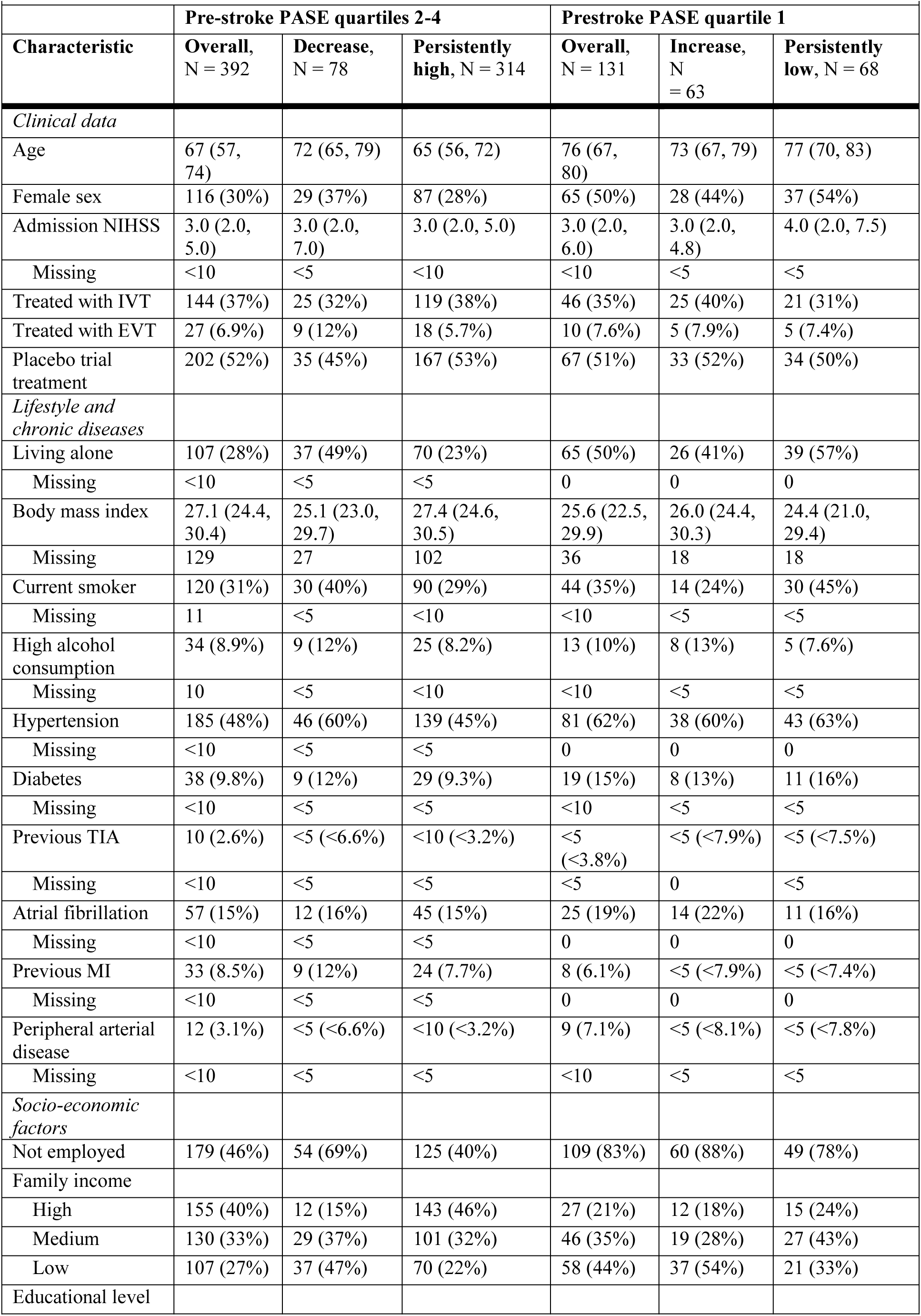
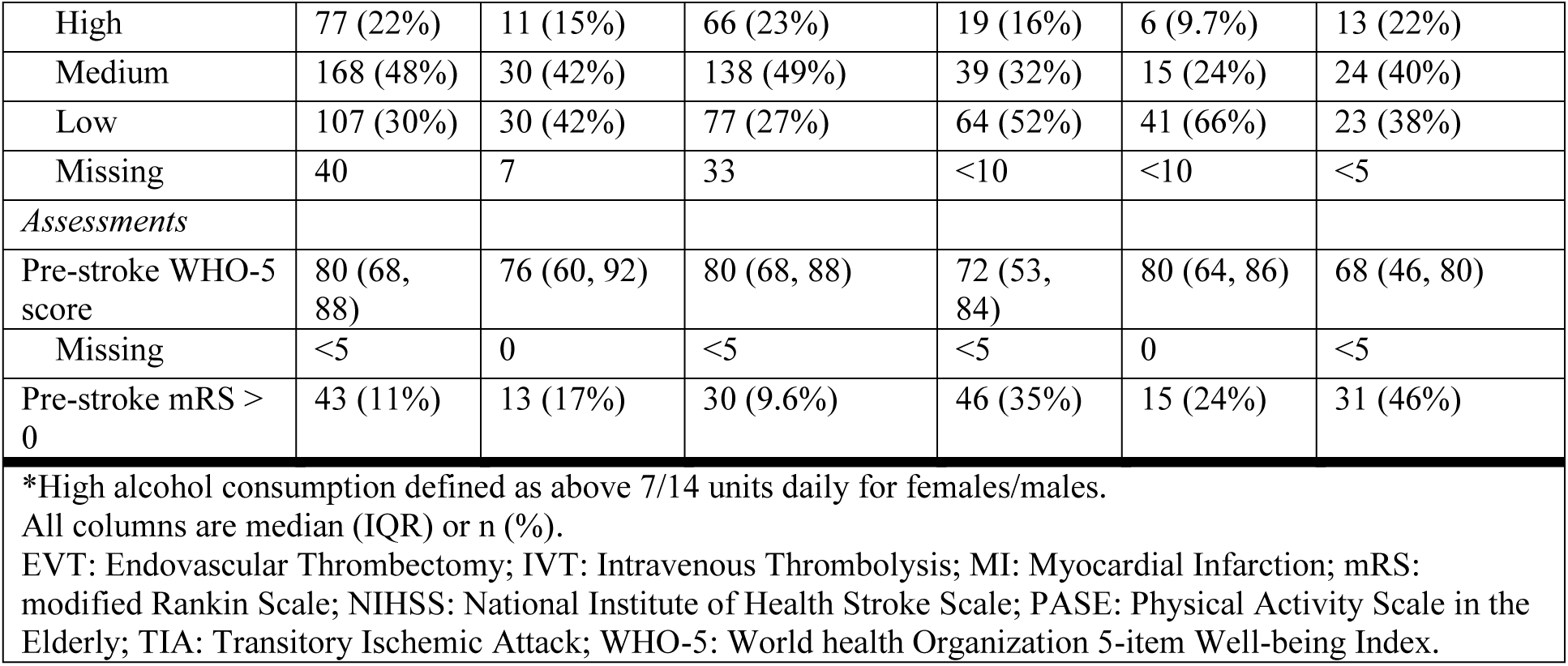
Baseline variables stratified by the direction of change in PA.

|  | Pre-stroke PASE quartiles 2-4 |  |  | Prestroke PASE quartile 1 |  |  |
| --- | --- | --- | --- | --- | --- | --- |
| Characteristic | Overall,<br>N = 392 | Decrease,<br>N = 78 | Persistently<br>high, N = 314 | Overall,<br>N = 131 | Increase,<br>N<br>= 63 | Persistently<br>low, N = 68 |
| *Clinical data* |  |  |  |  |  |  |
| Age | 67 (57, 74) | 72 (65, 79) | 65 (56, 72) | 76 (67, 80) | 73 (67, 79) | 77 (70, 83) |
| Female sex | 116 (30%) | 29 (37%) | 87 (28%) | 65 (50%) | 28 (44%) | 37 (54%) |
| Admission NIHSS | 3.0 (2.0, 5.0) | 3.0 (2.0, 7.0) | 3.0 (2.0, 5.0) | 3.0 (2.0, 6.0) | 3.0 (2.0, 4.8) | 4.0 (2.0, 7.5) |
| Missing | <10 | <5 | <10 | <10 | <5 | <5 |
| Treated with IVT | 144 (37%) | 25 (32%) | 119 (38%) | 46 (35%) | 25 (40%) | 21 (31%) |
| Treated with EVT | 27 (6.9%) | 9 (12%) | 18 (5.7%) | 10 (7.6%) | 5 (7.9%) | 5 (7.4%) |
| Placebo trial treatment | 202 (52%) | 35 (45%) | 167 (53%) | 67 (51%) | 33 (52%) | 34 (50%) |
| *Lifestyle and chronic diseases* |  |  |  |  |  |  |
| Living alone | 107 (28%) | 37 (49%) | 70 (23%) | 65 (50%) | 26 (41%) | 39 (57%) |
| Missing | <10 | <5 | <5 | 0 | 0 | 0 |
| Body mass index | 27.1 (24.4, 30.4) | 25.1 (23.0, 29.7) | 27.4 (24.6, 30.5) | 25.6 (22.5, 29.9) | 26.0 (24.4, 30.3) | 24.4 (21.0, 29.4) |
| Missing | 129 | 27 | 102 | 36 | 18 | 18 |
| Current smoker | 120 (31%) | 30 (40%) | 90 (29%) | 44 (35%) | 14 (24%) | 30 (45%) |
| Missing | 11 | <5 | <10 | <10 | <5 | <5 |
| High alcohol consumption | 34 (8.9%) | 9 (12%) | 25 (8.2%) | 13 (10%) | 8 (13%) | 5 (7.6%) |
| Missing | 10 | <5 | <10 | <10 | <5 | <5 |
| Hypertension | 185 (48%) | 46 (60%) | 139 (45%) | 81 (62%) | 38 (60%) | 43 (63%) |
| Missing | <10 | <5 | <5 | 0 | 0 | 0 |
| Diabetes | 38 (9.8%) | 9 (12%) | 29 (9.3%) | 19 (15%) | 8 (13%) | 11 (16%) |
| Missing | <10 | <5 | <5 | <10 | <5 | <5 |
| Previous TIA | 10 (2.6%) | <5 (<6.6%) | <10 (<3.2%) | <5 (<3.8%) | <5 (<7.9%) | <5 (<7.5%) |
| Missing | <10 | <5 | <5 | <5 | 0 | <5 |
| Atrial fibrillation | 57 (15%) | 12 (16%) | 45 (15%) | 25 (19%) | 14 (22%) | 11 (16%) |
| Missing | <10 | <5 | <5 | 0 | 0 | 0 |
| Previous MI | 33 (8.5%) | 9 (12%) | 24 (7.7%) | 8 (6.1%) | <5 (<7.9%) | <5 (<7.4%) |
| Missing | <10 | <5 | <5 | 0 | 0 | 0 |
| Peripheral arterial disease | 12 (3.1%) | <5 (<6.6%) | <10 (<3.2%) | 9 (7.1%) | <5 (<8.1%) | <5 (<7.8%) |
| Missing | <10 | <5 | <5 | <10 | <5 | <5 |
| *Socio-economic factors* |  |  |  |  |  |  |
| Not employed | 179 (46%) | 54 (69%) | 125 (40%) | 109 (83%) | 60 (88%) | 49 (78%) |
| Family income |  |  |  |  |  |  |
| High | 155 (40%) | 12 (15%) | 143 (46%) | 27 (21%) | 12 (18%) | 15 (24%) |
| Medium | 130 (33%) | 29 (37%) | 101 (32%) | 46 (35%) | 19 (28%) | 27 (43%) |
| Low | 107 (27%) | 37 (47%) | 70 (22%) | 58 (44%) | 37 (54%) | 21 (33%) |
| Educational level |  |  |  |  |  |  |
| High | 77 (22%) | 11 (15%) | 66 (23%) | 19 (16%) | 6 (9.7%) | 13 (22%) |
| Medium | 168 (48%) | 30 (42%) | 138 (49%) | 39 (32%) | 15 (24%) | 24 (40%) |
| Low | 107 (30%) | 30 (42%) | 77 (27%) | 64 (52%) | 41 (66%) | 23 (38%) |
| Missing | 40 | 7 | 33 | <10 | <10 | <5 |
| *Assessments* |  |  |  |  |  |  |
| Pre-stroke WHO-5 score | 80 (68, 88) | 76 (60, 92) | 80 (68, 88) | 72 (53, 84) | 80 (64, 86) | 68 (46, 80) |
| Missing | <5 | 0 | <5 | <5 | 0 | <5 |
| Pre-stroke mRS > 0 | 43 (11%) | 13 (17%) | 30 (9.6%) | 46 (35%) | 15 (24%) | 31 (46%) |
| *High alcohol consumption defined as above 7/14 units daily for females/males.<br>All columns are median (IQR) or n (%).<br>EVT: Endovascular Thrombectomy; IVT: Intravenous Thrombolysis; MI: Myocardial Infarction; mRS: modified Rankin Scale; NIHSS: National Institute of Health Stroke Scale; PASE: Physical Activity Scale in the Elderly; TIA: Transitory Ischemic Attack; WHO-5: World health Organization 5-item Well-being Index. |  |  |  |  |  |  |

A total of 78 (19.9 %) patients experienced a decrease in PA to the 1^st^ PASE quartile at six months as compared to their pre-stroke PA level. Of the patients in the 1^st^ PASE quartile pre-stroke, 63 (48.1 %) patients had increased their PA level at six months. In Figure 1 PA level changes are visualized.

**Figure 1.**
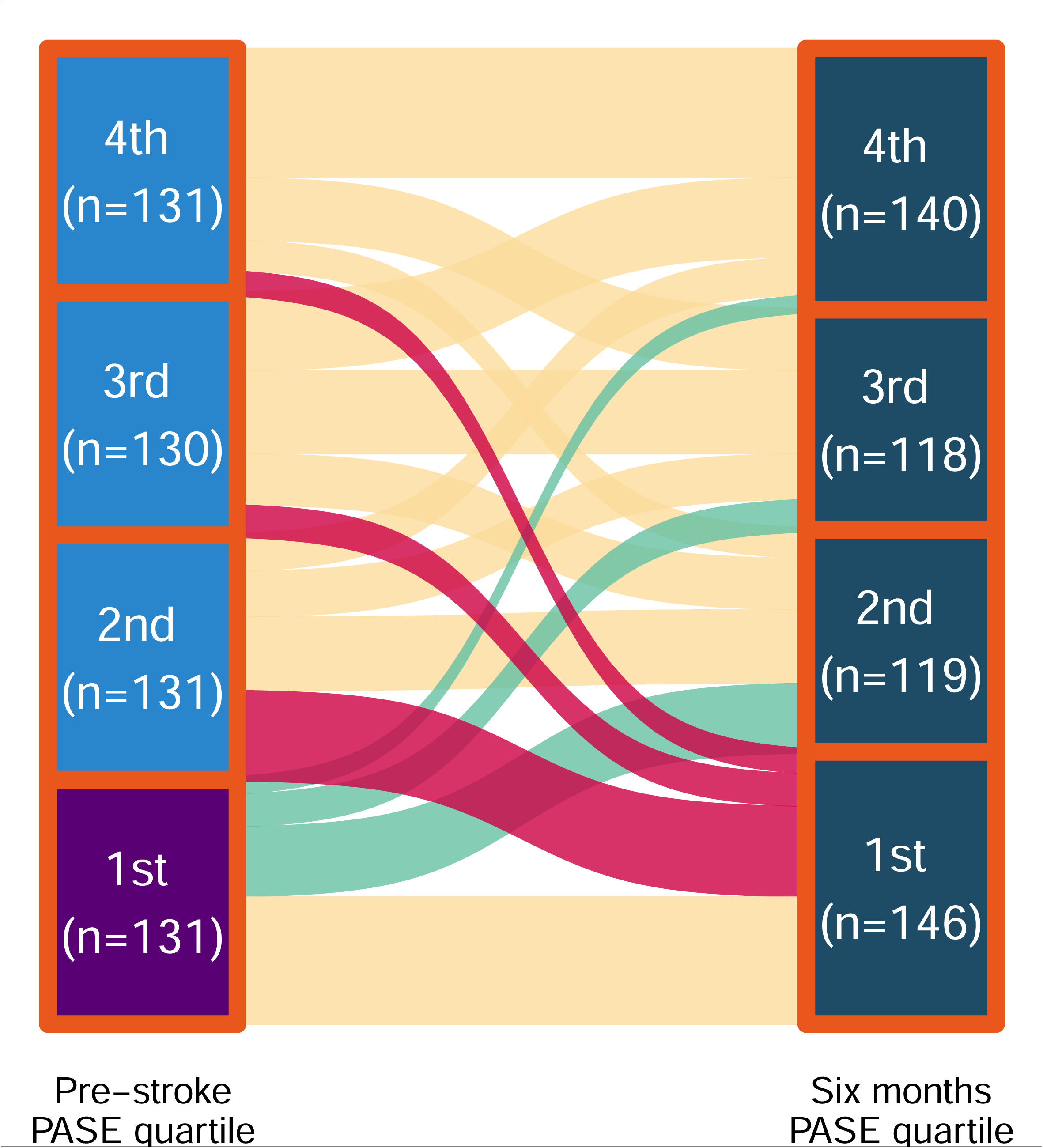
PA level changes from pre-stroke to post-stroke. Marked in green are patients with increased level of PA from low to a higher level. Red marks patients with decreased level of PA from a higher level to a low level.

Between sexes, the PA groups distribution differed as females reported lower PASE scores both prior to stroke and at follow-up. Females were also older, had a higher NIHSS score, more often lived alone, more were not employed, had a lower household income and had a lower educational level. Males reported higher alcohol consumption and more had diabetes. Please refer to Supplementary table 1 for all details.

Patients excluded, including patients who died, from the TALOS cohort were significantly older, had higher NIHSS and lower BMI, were less often employed and had lower pre-stroke WHO-5 scores (data not shown).^25^

### Regression analyses

In the multivariable linear regression analysis, we found that higher age (-0.86; CI: -1.8, - 0.08; per year), higher acute NIHSS score (-2.3; CI: -4.2, -0.39; per point) and low family income (-30; CI: -50, -10) correlated to a significantly lower six months PASE score. Variables that correlated to a higher score were higher pre-stroke PASE score (0.32, CI: 0.22, 0.42; per point), atrial fibrillation (22, CI: 2.8;40) and higher WHO-5 well-being score (0.60; CI: 0.26, 0.95; per point). Please see Table 2.

**Table 2:**
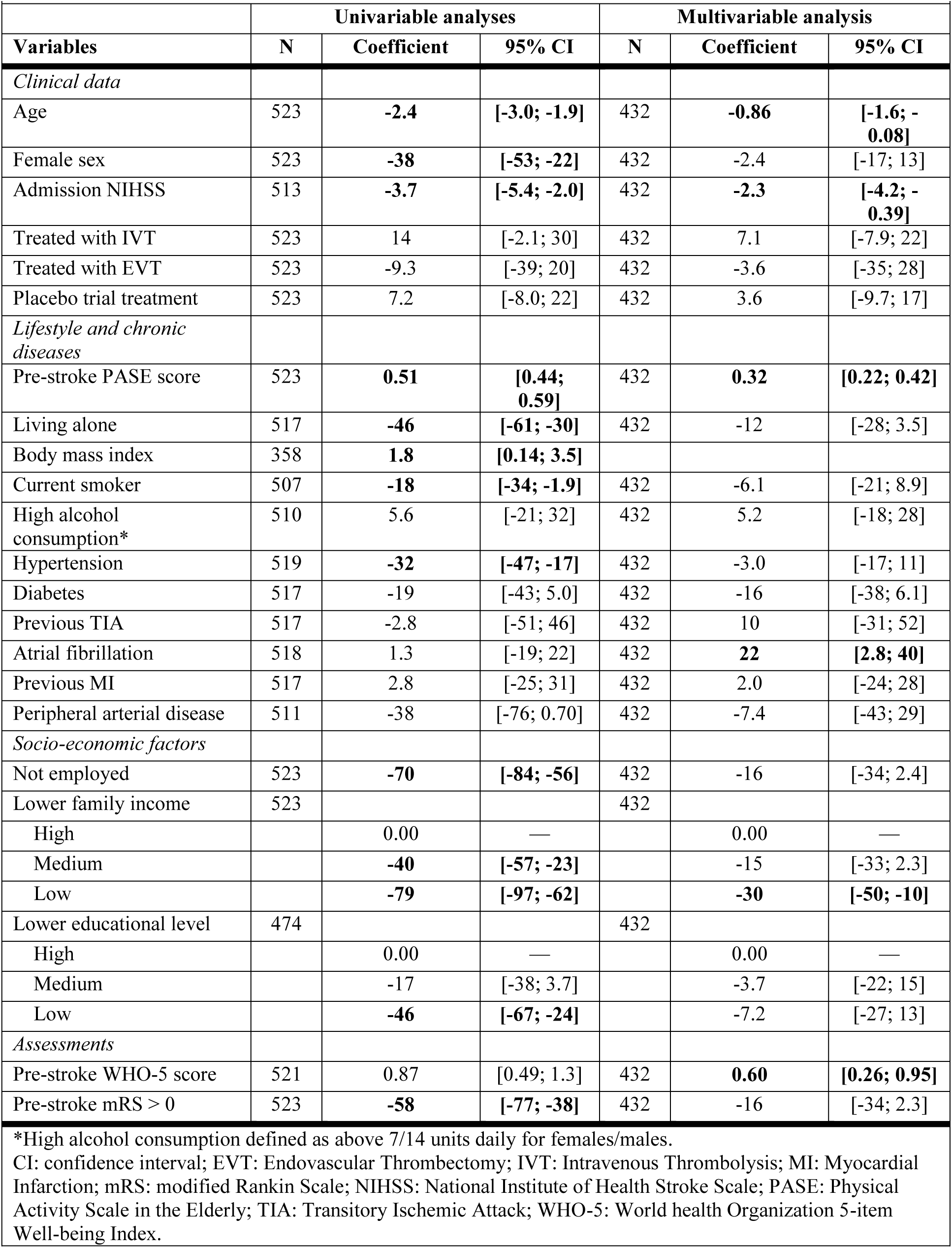
Linear regression analysis of six months PASE score.

| Table 2: Linear regression analysis of six months PASE score |  |  |  |  |  |  |
| --- | --- | --- | --- | --- | --- | --- |
|  | Univariable analyses |  |  | Multivariable analysis |  |  |
| Variables | N | Coefficient | 95% CI | N | Coefficient | 95% CI |
| *Clinical data* |  |  |  |  |  |  |
| Age | 523 | **-2.4** | **[-3.0; -1.9]** | 432 | **-0.86** | **[-1.6; -0.08]** |
| Female sex | 523 | **-38** | **[-53; -22]** | 432 | -2.4 | [-17; 13] |
| Admission NIHSS | 513 | **-3.7** | **[-5.4; -2.0]** | 432 | **-2.3** | **[-4.2; -0.39]** |
| Treated with IVT | 523 | 14 | [-2.1; 30] | 432 | 7.1 | [-7.9; 22] |
| Treated with EVT | 523 | -9.3 | [-39; 20] | 432 | -3.6 | [-35; 28] |
| Placebo trial treatment | 523 | 7.2 | [-8.0; 22] | 432 | 3.6 | [-9.7; 17] |
| *Lifestyle and chronic diseases* |  |  |  |  |  |  |
| Pre-stroke PASE score | 523 | **0.51** | **[0.44; 0.59]** | 432 | **0.32** | **[0.22; 0.42]** |
| Living alone | 517 | **-46** | **[-61; -30]** | 432 | -12 | [-28; 3.5] |
| Body mass index | 358 | **1.8** | **[0.14; 3.5]** |  |  |  |
| Current smoker | 507 | **-18** | **[-34; -1.9]** | 432 | -6.1 | [-21; 8.9] |
| High alcohol consumption* | 510 | 5.6 | [-21; 32] | 432 | 5.2 | [-18; 28] |
| Hypertension | 519 | **-32** | **[-47; -17]** | 432 | -3.0 | [-17; 11] |
| Diabetes | 517 | -19 | [-43; 5.0] | 432 | -16 | [-38; 6.1] |
| Previous TIA | 517 | -2.8 | [-51; 46] | 432 | 10 | [-31; 52] |
| Atrial fibrillation | 518 | 1.3 | [-19; 22] | 432 | **22** | **[2.8; 40]** |
| Previous MI | 517 | 2.8 | [-25; 31] | 432 | 2.0 | [-24; 28] |
| Peripheral arterial disease | 511 | -38 | [-76; 0.70] | 432 | -7.4 | [-43; 29] |
| *Socio-economic factors* |  |  |  |  |  |  |
| Not employed | 523 | **-70** | **[-84; -56]** | 432 | -16 | [-34; 2.4] |
| Lower family income | 523 |  |  | 432 |  |  |
| High |  | 0.00 | — |  | 0.00 | — |
| Medium |  | **-40** | **[-57; -23]** |  | -15 | [-33; 2.3] |
| Low |  | **-79** | **[-97; -62]** |  | **-30** | **[-50; -10]** |
| Lower educational level | 474 |  |  | 432 |  |  |
| High |  | 0.00 | — |  | 0.00 | — |
| Medium |  | -17 | [-38; 3.7] |  | -3.7 | [-22; 15] |
| Low |  | **-46** | **[-67; -24]** |  | -7.2 | [-27; 13] |
| *Assessments* |  |  |  |  |  |  |
| Pre-stroke WHO-5 score | 521 | 0.87 | [0.49; 1.3] | 432 | **0.60** | **[0.26; 0.95]** |
| Pre-stroke mRS > 0 | 523 | **-58** | **[-77; -38]** | 432 | -16 | [-34; 2.3] |
| *High alcohol consumption defined as above 7/14 units daily for females/males.<br>CI: confidence interval; EVT: Endovascular Thrombectomy; IVT: Intravenous Thrombolysis; MI: Myocardial Infarction; mRS: modified Rankin Scale; NIHSS: National Institute of Health Stroke Scale; PASE: Physical Activity Scale in the Elderly; TIA: Transitory Ischemic Attack; WHO-5: World health Organization 5-item Well-being Index. |  |  |  |  |  |  |

### Predictors for a decrease in PA

The model predicting decrease in PA level had a sensitivity of 0.887, a specificity of 0.331, a positive predictive value (PPV) of 0.673 and a negative predictive value (NPV) of 0.654. The median AUROC was 0.675. The most important variables predicting a decreased PA were living alone (OR 2.4), EVT treatment (OR 1.9), lower family income (OR 1.6), hypertension (OR 1.4) and no employment (OR 1.4).

Median tuning parameters for the pooled models were alpha=1 and lambda=0.013.

### Predictors for an increase in PA

The model predicting increase in PA level performed with a sensitivity of 0.618 and specificity of 0.575, PPV was 0.617 and NPV was 0.576, while the median AUROC was 0.619. The most important variables affecting prediction of PA increase were current smoking (OR 0.55), lower educational level (OR 0.59), previous TIA (OR 0.62), pre-stroke mRS > 0 (0.74) and no employment (OR 0.79).

Median tuning parameters were alpha=1 and lambda=0.038.

Please refer to Figure 2 for calibration plots as well as Table 3 and Figure 3 for an overview of all variables in both models.

**Figure 2.**
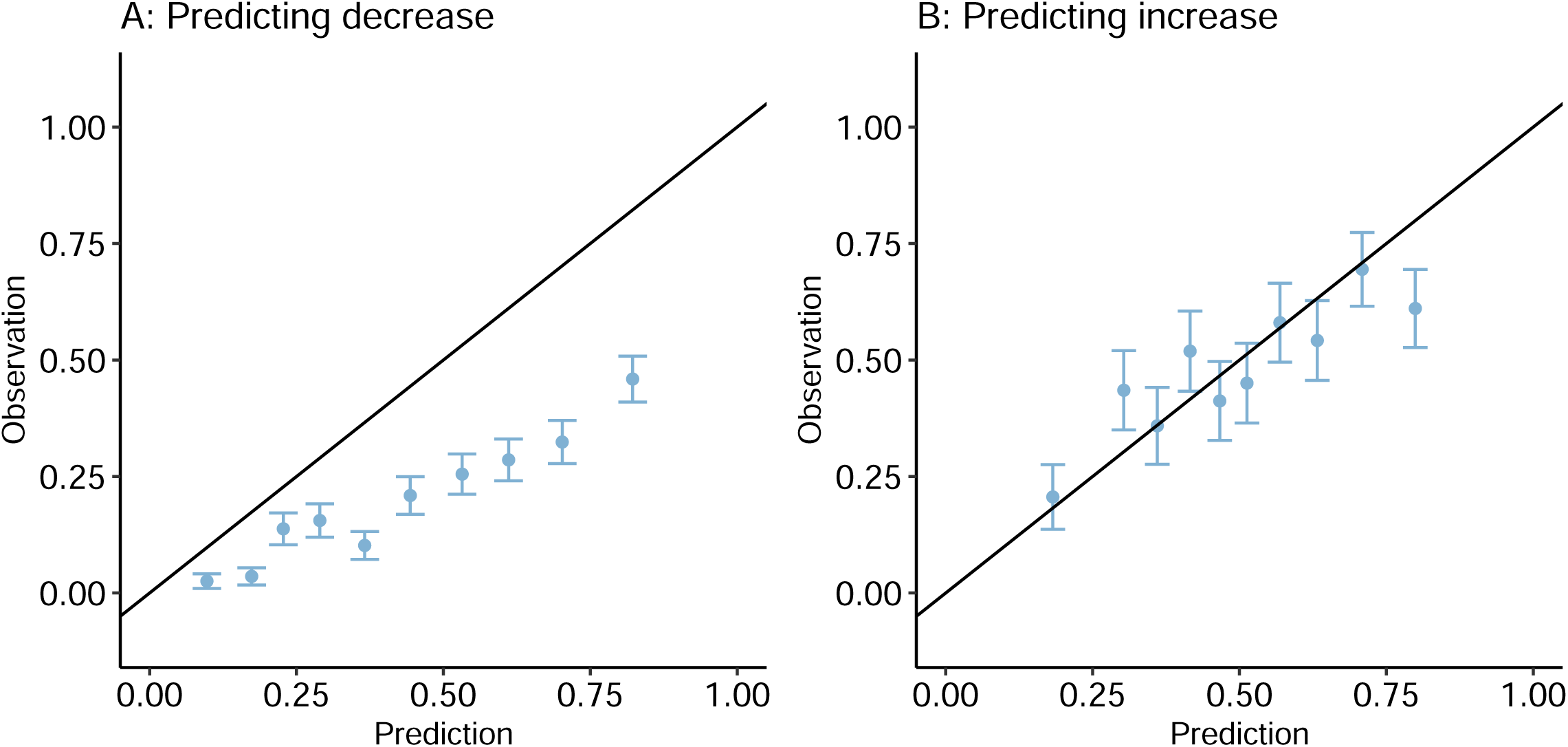
Calibration plots. Calibration plots of the two prediction models. A calibration plot compares the model prediction probability to the fraction of observed events in the data set in binned groups (here each 10 %). In a well calibrated model, the predicted probability and fraction of observed events should be proportional with a slope of one (i.e. follow the plotted black line).

**Figure 3.**
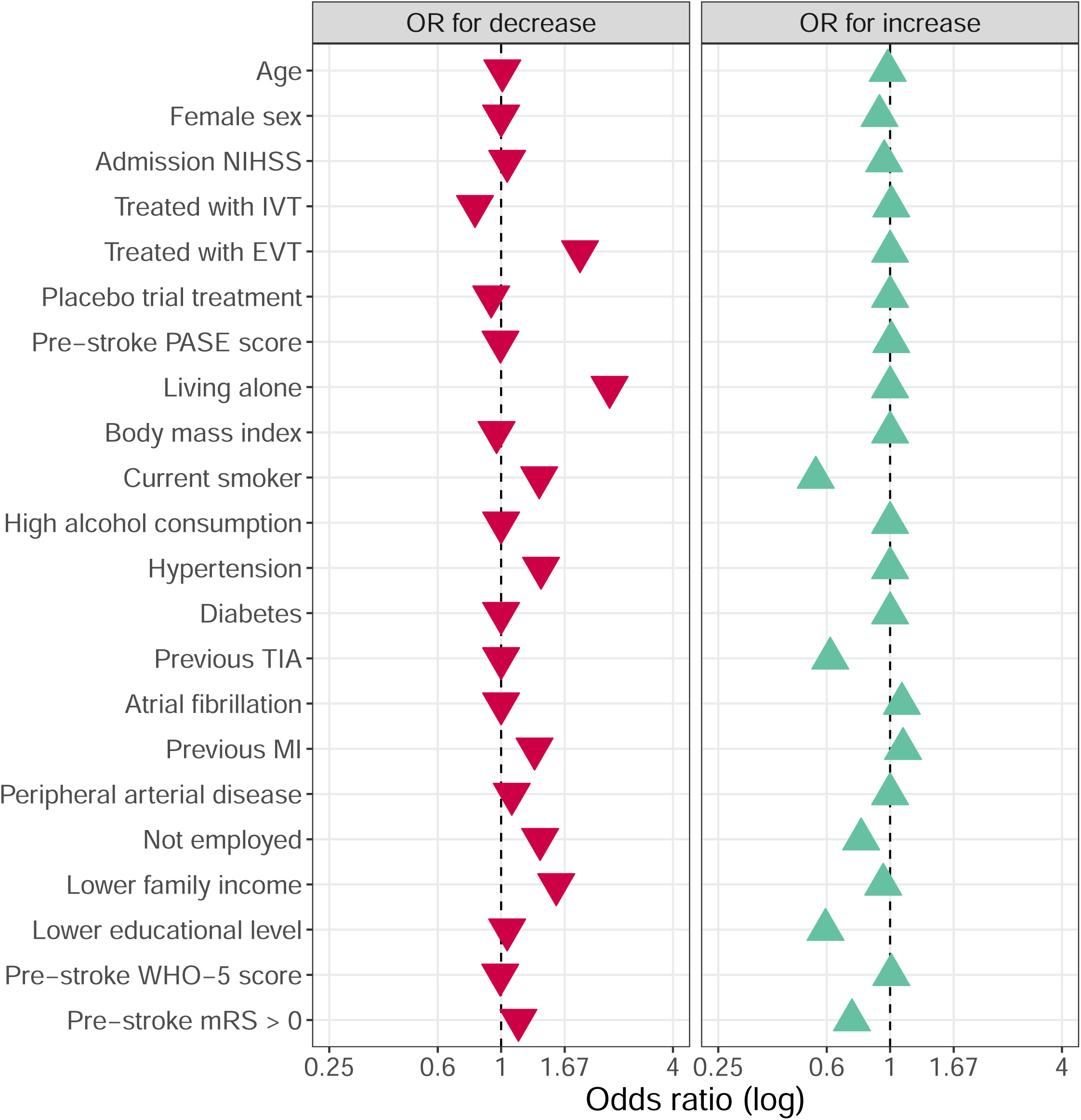
Visualization of odds ratios for each prediction models. High alcohol consumption is defined as above 7/14 units daily for females/males. EVT: Endovascular Thrombectomy; IVT: Intravenous Thrombolysis; MI: Myocardial Infarction; mRS: modified Rankin Scale; NIHSS: National Institute of Health Stroke Scale; PASE: Physical Activity Scale in the Elderly; TIA: Transitory Ischemic Attack; WHO-5: World health Organization 5-item Well-being Index.

**Table 3:** Combined table of model ORs from the two prediction models.

| Table 3: Combined table of model ORs from the two prediction models |  |  |  |  |
| --- | --- | --- | --- | --- |
|  | Decrease to low PA level |  | Increase to higher PA level |  |
| Variables | Median OR | Mean OR | Median OR | Mean OR |
| *Clinical data* |  |  |  |  |
| Age | **1.01** | **1.01** | **0.981** | **0.980** |
| Female sex | 1.00 | 0.979 | **0.918** | **0.876** |
| Admission NIHSS | **1.05** | **1.05** | **0.954** | **0.955** |
| Treated with IVT | **0.810** | **0.825** | **1.01** | **1.08** |
| Treated with EVT | **1.89** | **1.86** | 1.00 | 1.01 |
| Placebo trial treatment | **0.923** | **0.883** | 1.00 | 1.01 |
| *Lifestyle and chronic diseases* |  |  |  |  |
| Pre-stroke PASE score | **0.995** | **0.995** | **1.01** | **1.01** |
| Living alone | **2.40** | **2.41** | 1.00 | 0.924 |
| Body mass index | **0.964** | **0.964** | 1.00 | 1.01 |
| Current smoker | **1.36** | **1.35** | **0.549** | **0.545** |
| High alcohol consumption* | 1.00 | 1.04 | 1.00 | 1.13 |
| Hypertension | **1.38** | **1.39** | 1.00 | 0.959 |
| Diabetes | 1.00 | 1.01 | 1.00 | 0.919 |
| Previous TIA | 1.00 | 0.909 | **0.617** | **0.537** |
| Atrial fibrillation | 1.00 | 0.969 | **1.10** | **1.15** |
| Previous MI | **1.31** | **1.31** | **1.11** | **1.30** |
| Peripheral arterial disease | **1.09** | **1.18** | 1.00 | 0.970 |
| *Socio-economic factors* |  |  |  |  |
| Not employed | **1.37** | **1.36** | **0.792** | **0.795** |
| Lower family income | **1.56** | **1.55** | **0.946** | **0.924** |
| Lower educational level | **1.05** | **1.07** | **0.594** | **0.596** |
| *Assessments* |  |  |  |  |
| Pre-stroke WHO-5 score | **0.993** | **0.993** | **1.01** | **1.01** |
| Pre-stroke mRS > 0 | **1.15** | **1.15** | **0.735** | **0.747** |
| *High alcohol consumption defined as above 7/14 units daily for females/males.<br>EVT: Endovascular Thrombectomy; IVT: Intravenous Thrombolysis; MI: Myocardial Infarction; mRS: modified Rankin Scale; NIHSS: National Institute of Health Stroke Scale; PASE: Physical Activity Scale in the Elderly; TIA: Transitory Ischemic Attack; WHO-5: World health Organization 5-item Well-being Index.<br>Variable ORs where both median and mean values are $\neq 1$ are considered statistically relevant and highlighted in bold. |  |  |  |  |

## Discussion

Among 523 first-ever ischemic stroke patients, we found that approximately one in five patients experienced a decrease in PA six months after stroke to the lowest quartile. But also, that approximately half of patients who were in the lowest quartile prior to stroke, increased their PA level to a higher quartile six months after stroke.

In the multivariable linear regression model, we found that higher age, higher NIHSS score at stroke admission, and lower family income level were associated with a lower PASE score six months after stroke, and higher pre-stroke PASE score, atrial fibrillation and higher WHO-5 well-being score were associated to a higher score. Notably, a higher pre-stroke PASE score was correlated to higher post-stroke score by only 0.32 points per point, indicating that other factors besides habits influence PA level. Also, known or newly diagnosed atrial fibrillation was associated to higher six months PASE score, which may reflect, that these patients receive focused intervention as part of their cardiological evaluation.

We developed two machine-learning models to predict changes in PA based on two clinically different patient groups among first ever stroke patients: patients in the three highest PASE quartiles and patients in the lowest PASE quartile group. The overall model performance was only modest in both cases, but the statistical model used allows for some inference on the variables included.

In predicting decrease, the model showed that living alone, endovascular treatment (signifying a more severe stroke at onset), lower family income, no employment, and hypertension were the most important factors, all with higher odds for a decrease in PA level. The model showed a high sensitivity, but poor calibration. In predicting increase from the lowest PA level to a higher level, smoking, lower educational level, previous TIA diagnosis, pre-stroke mRS > 0, and no employment, all showed lower odds of increase. The findings are in line with previous studies indicating a positive correlation between SES and health status and PA, though patterns of PA may be different in SES groups as lower paid work tend to be more physically demanding, while people in higher SES groups tend to spend more time on PA during leisure time.^40,41^

Only two other previous studies were identified to have reported PA both before and after stroke. Botö et al,^22^ found that pre-stroke physical inactivity and stroke severity were associated with post-stroke physical inactivity. They included 190 patients with first ever and recurrent stroke as well as both ischemic and hemorrhagic strokes, representing a very heterogeneous stroke population. Stroke severity, fear of falling and pre-stroke physical inactivity were associated to post-stroke inactivity. Unfortunately, 43% of the study population was lost to follow-up which increases the risk of selection bias. The present study has a larger sample size (n=523) and a low number of lost to follow-up and uses a more detailed and concrete questionnaire taking many different every-day and household activities into account when calculating a continuous score for a more nuanced measure on PA.

The other study was the Danish ExStroke trial by Krarup et al, which also used the PASE questionnaire.^20^ This was a randomized trial on PA intervention including patients with ischemic stroke within 3 months after stroke, that found no effect of verbal intervention to change PA levels after 6 months. The median PASE in the ExStroke trial was 70 but was recorded at a median of 10 [5;23,5] days after stroke onset, which may explain the difference from this current trial. The ExStroke trial also reported no significant change in in overall PASE score at 6 months, but they did not report any data on patient-level change.

There are important limitations to the present study. It is a post-hoc analysis of patients included in the TALOS trial, and the population constitutes primarily of patients with mild ischemic strokes (median NIHSS was 3), which may limit the generalizability of the results. The PASE score includes time spent at work independently of the type of work and was developed for people aged 65+ resulting in working contributing with a relatively high score compared to other activities measured by the scale. The analyses are, however, adjusted for employment, which should counteract this imbalance. The PASE score may also have other limitations such as only regarding the seven days prior to questioning and generalizing this measure, but has been shown valid as a measure of physical activity.^27^

Strengths of this study includes, that patients were included with no age limit and pre-stroke PA was assessed shortly after stroke admission. Patients were included by consent from next of kin if not able themselves, which have extended the possible range of acute stroke symptoms, though we did not have PA information from the most severely disabled patients and those lost to follow-up.^25^ Patients included were all first-ever stroke patients, who are expected to be more homogeneous in their post-stroke behavior. The data included were comprehensive and reflects the data available upon stroke admission, that would generally be available in a clinical setting.^28^

Several factors may limit the predictive performance of our models. The model to predict decrease showed signs of overfitting to the data with high sensitivity and low specificity as well as problematic calibration. This may have been caused by the pre-stroke high PA level group being more heterogeneous compared to the low pre-stroke PA group. Other quantitative biological measures not available for inclusion in these models could be retrieved from regular laboratory tests or imaging such as infarct location and volume, or small vessel disease burden to potentially increase model performance. Qualitative measures on social life or feeling of loneliness, may also be relevant, but difficult to incorporate in a statistical model.^42^ No specific steps were taken to ensure demographic diversity in the patient population or in the trial steering committee.

In conclusion, change in PA level after first-time ischemic stroke is highly dynamic. This study shows that positive change is possible, as half of patients in the lowest PA quartile increased to a higher level after stroke. Awareness, however, should also be paid to the 20% of patients decreasing to the lowest PA quartile as well as to the patients with a persistently low PA level. Despite comprehensive data, predicting change in PA was only possible with modest results but showed that socioeconomic status and lifestyle factors were important for PA change. Our study should encourage further research into the underlying factors at play in physical activity behavior after stroke, and guide future rehabilitation efforts and help identify vulnerable patients to prevent inactivity or improve PA after stroke.

## Non-standard Abbreviations and Acronyms

PA: Physical Activity
PASE: Physical Activity Scale for the Elderly
SES: Socioeconomic status
TALOS: The Efficacy of Citalopram Treatment in Acute Ischemic Stroke
IQR: Interquartile range
NIHSS: National Institute of Health Stroke Scale
WHO-5: World Health Organization 5-item Well-being Index
mRS: modified Rankin Scale
FED: Family equivalent disposable
CI: confidence intervals
BMI: body mass index
OR: Odds ratio
AUROC: Area under receiver operated curve
PPV: Positive predictive value
NPV: Negative predictive value

## Acknowledgments

None

## Supplemental Material

Tables S1

